# To what extent do we need to rely on non-pharmaceutical interventions while COVID-19 vaccines roll out in 2021?

**DOI:** 10.1101/2021.02.03.21251108

**Authors:** Juan Yang, Valentina Marziano, Xiaowei Deng, Giorgio Guzzetta, Juanjuan Zhang, Filippo Trentini, Jun Cai, Piero Poletti, Wen Zheng, Wei Wang, Qianhui Wu, Zeyao Zhao, Kaige Dong, Guangjie Zhong, Cécile Viboud, Stefano Merler, Marco Ajelli, Hongjie Yu

## Abstract

COVID-19 vaccination is being conducted in over 190 countries/regions to control SARS-CoV-2 transmission and return to a pre-pandemic lifestyle. However, understanding when non-pharmaceutical interventions (NPIs) can be lifted as immunity builds up remain a key question for policy makers. To address it, we built a data-driven model of SARS-CoV-2 transmission for China. We estimated that to prevent the escalation of local outbreaks to widespread epidemics, stringent NPIs need to remain in place at least one year after the start of vaccination. Should NPIs alone be capable to keep the reproduction number (R_t_) around 1.3, the synergetic effect of NPIs and vaccination could reduce up to 99% of COVID-19 burden and bring R_t_ below the epidemic threshold in about 9 months. Maintaining strict NPIs throughout 2021 is of paramount importance to reduce COVID-19 burden while vaccines are distributed to the population, especially in large populations with little natural immunity.

---

The novel coronavirus disease 2019 (COVID-19) pandemic is far from over with cases still surging in many countries across the globe, particularly with India suffering from a catastrophic second wave ^1^. In 2020, epidemic suppression and/or mitigation have relied on non-pharmaceutical interventions (NPIs), including social distancing, school closure, masking, and case isolation. Although effective and widely adopted to limit SARS-CoV-2 transmission and reduce COVID-19 burden, these interventions entail enormous economic costs and negatively affect quality of life ^2^. Additionally, in many countries, relaxation of NPIs has led to a resurgence of the epidemic as herd immunity has not been reached thus far ^3^.

Effective vaccines against COVID-19 remain the only foreseeable means of both suppressing the infection and returning to pre-pandemic social and economic activity patterns. Globally, several vaccines have been licensed, and vaccination programs have been initiated in more than 190 countries/regions, including China ^4^. However, the projected global production and delivery capacities are likely to be inadequate to provide COVID-19 vaccines to all individuals who are still susceptible to SARS-CoV-2 infection ^3^. The effectiveness of COVID-19 vaccination campaigns will depend on several factors, including pre-existing immunity, vaccine supply, willingness to receive the vaccine, and strategies for vaccine allocation and deployment ^5^.

To avoid widespread transmission of SARS-CoV-2, since the end of the first COVID-19 wave in the spring of 2020, China has implemented strict NPIs and has successfully controlled local outbreaks, preventing a second widespread wave of COVID-19. Since December 2020, China has given conditional approval for the use of four COVID-19 vaccines and emergency use approval for one. As of May 7, 2021, 308 million doses (corresponding to 10.7% of the population) have been administered^6^. However, such a coverage is still extremely low and thus China remains highly vulnerable to importations of SARS-CoV-2 and onward transmission, as proved by several local outbreaks that occurred in the first four months of 2021, the largest of which occurring in Heilongjiang province led to 659 reported cases and spilled over to a neighboring province (419 cases were reported in Jilin) ^7^. At present, estimating whether and when NPIs can be lifted, and the extent to which we need to rely on NPIs while vaccines roll out represents a top priority for policy making.

This question has not been well addressed in China, one of the few countries in the world where nearly the entire population is still susceptible to SARS-CoV-2 infection, and home to almost 1.4 billion individuals (roughly 18% of the world population). To fill this gap, we built on top of the wide body of work adopting mathematical models of the infection transmission process to evaluate vaccination programs ^8–12^. In particular, we developed an age-structured stochastic model to simulate SARS-CoV-2 transmission triggered by cases imported in mainland China, based on a susceptible-infectious-removed (SIR) scheme (Supplementary Fig.1). We consider a situation in which: i) no ongoing widespread SARS-CoV-2 transmission, ii) nearly no immunity in the population, and iii) high risk of importing SARS-CoV-2 infected individuals, possibly leading to an upsurge of COVID-19 cases. Since COVID-19 vaccines are expected to continue rolling out throughout 2021-2022, we consider also alternative scenarios where SARS-CoV-2 infections leading to an outbreak are imported when 10% (close to the coverage as of May 2021), 20%, and 30% of the Chinese population has already been vaccinated (according to the simulated vaccination program).

In the model, we account for heterogeneous mixing patterns by age ^13^ and progressive vaccine deployment among different population segments based on a priority scheme (essential workers, older adults and individuals with underlying conditions, etc.) ^14^.

Further, we overlay a disease burden model on the transmission model to estimate the number of symptomatic cases, hospitalizations, ICU admissions, and deaths under different vaccination scenarios and based on empirical data ^15–20^. The resulting integrated model is informed by data on COVID-19 natural history, age-mixing patterns specific to China quantified during the pre-pandemic period, and the size of the different vaccination targets in the Chinese population (e.g., individuals with pre-existing conditions). A qualitative model description is reported in the Methods section, a summary of model parameters and data sources is reported in Supplementary Table 1, all other details are reported in Supplementary File 1-5.

The combined effect of NPIs and vaccination programs are evaluated in terms of their ability in reducing the disease burden caused by outbreaks arising from possible importation of cases.

We considered a baseline vaccination scenario where: 1) vaccination starts 15 days after an outbreak triggered by 40 breakthrough imported SARS-CoV-2 infections; 2) vaccine efficacy (VE) against SARS-CoV-2 infections for a two-dose schedule (with a 21-day interval) is set at 80% ^21^; 3) vaccination coverage is capped at 70% across all ages ^14^; 4) 6 million doses are administered daily (4 per 1,000 individuals, informed by the ongoing COVID-19 vaccination program ^6^, and estimates of vaccine supply till 2021 in China ^22–26^); 5) the first priority target consists of older adults and individuals with underlying conditions (descriptions in details shown in Supplementary Table 2); 6) there is no prior population immunity from natural infection, which aligns with the situation in most of China where there has been little circulation of SARS-CoV-2 as of May 2021 ^3^; 7) we assume an initial reproductive number R_t_ =2.5 at the start of the outbreak ^27–32^, in the absence of NPIs and vaccination; 8) Children under 15 years of age were considered to have a lower susceptibility to SARS-CoV-2 infection as compared to adults (i.e., individuals aged 15 to 64 years), while individuals aged 65+ years had the highest susceptibility to infection (Supplementary Table 1) ^33, 34^; and 9) we let the model run for two years. To evaluate the impact of the baseline assumptions on our results, we conduct comprehensive sensitivity analyses (SA).

## Results

### Main analysis

In the absence of NPIs, the vaccination program is too slow to lower and delay the epidemic (Fig. 1a) and does not effectively reduce COVID-19 burden. R_t_ falls below the epidemic threshold (<1) 69 days after the epidemic start (Fig. 1b), but this is primarily attributable to immunity gained through natural infection rather than vaccination. Indeed, in this time frame, 52.2% of population gets infected, while only 6.7% of population has been vaccinated (Fig. 1c). The cumulative number of symptomatic cases and deaths over a 2-year period only decrease by 3.3% (95%CI, 3.1%-4.7%) and 6.7% (95%CI, 4.5%-8.9%), respectively, as compared to a reference scenario where there is no vaccination and no NPIs, which would lead to 306.73 million (95%CI, 282.68-320.60) symptomatic cases, 99.25 million (95%CI, 92.55-104.51) hospitalizations, 7.19 million (95%CI, 6.00-7.83) ICU admissions, and 9.38 million (95%CI, 7.70-10.26) deaths (Fig. 2).

**Figure 1.**
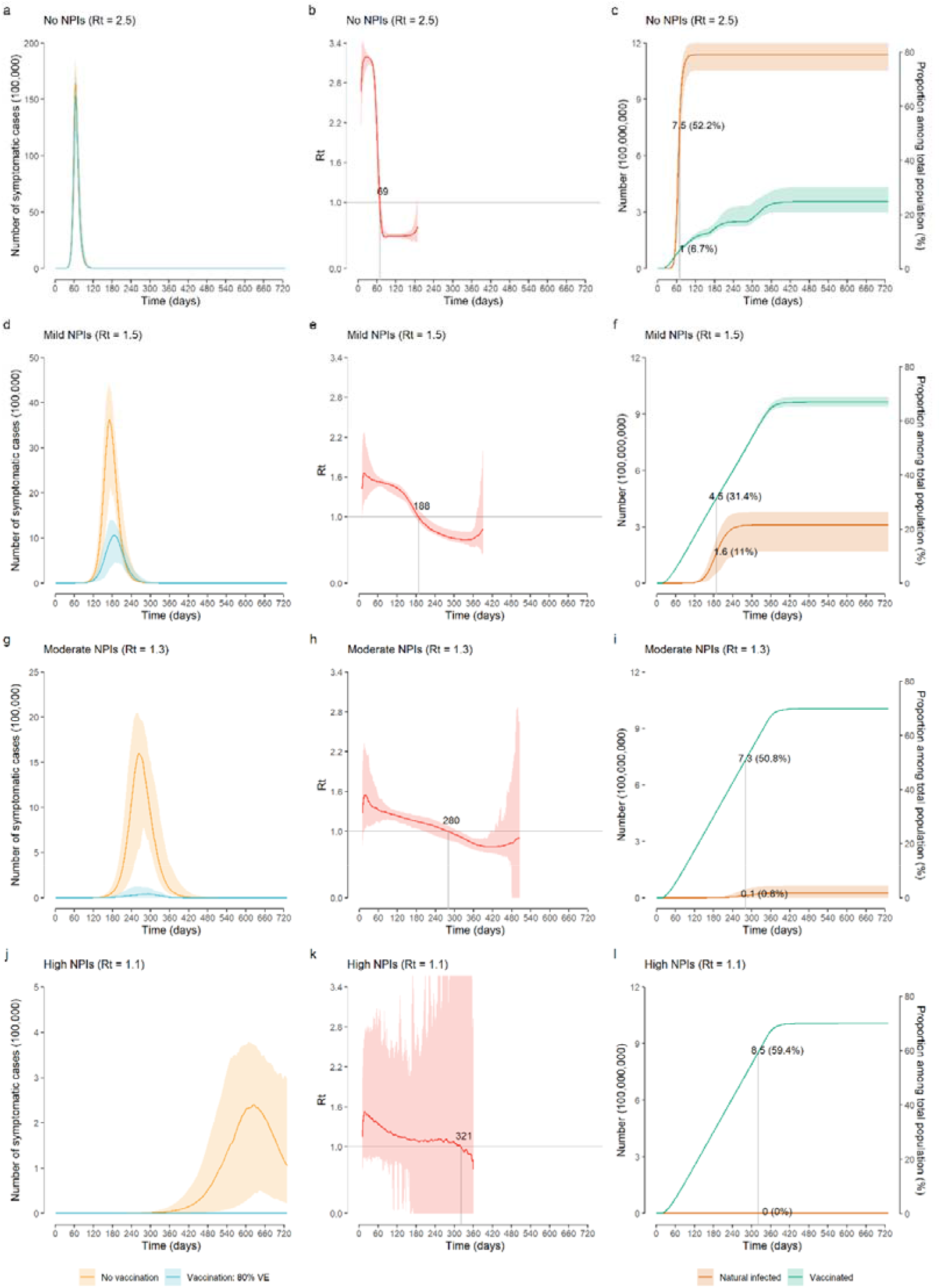
Time series of symptomatic cases, effective reproductive number R_t_, and population infected and vaccinated. a) Number of symptomatic cases over time as estimated in the no-NPIs scenario (initial R_t_=2.5) in the absence/presence of vaccination. b) Net reproduction number R_t_ over time, as estimated using a Bayesian framework (Supplementary File 6) from the time series of symptomatic cases in the no-NPIs scenario in the presence of vaccination. The horizontal line indicates the epidemic threshold R_t_=1 and the vertical line indicates where R_t_ cross this threshold. Note that for the first few generations of cases, R_t_ shows an increasing pattern linked to the highly stochastic nature of epidemics in their initial phase when epidemics with initially larger R_t_ are more likely to survive ^56^. c) Absolute numbers and proportion of the Chinese population infected and vaccinated over time in the no-NPIs scenario in the presence of vaccination. The population size of China in 2020 is 1,439,324,000 ^57^. d)-f): as a-c but for the mild NPIs scenario (initial R_t_=1.5). g)-i): as a-c but for the moderate NPIs scenario (initial R_t_=1.3); j)-l): As a-c but for the high NPIs scenario (initial R_t_=1.1). Line denotes median, and shadow denotes quantiles 0.025 and 0.975.

**Figure 2.**
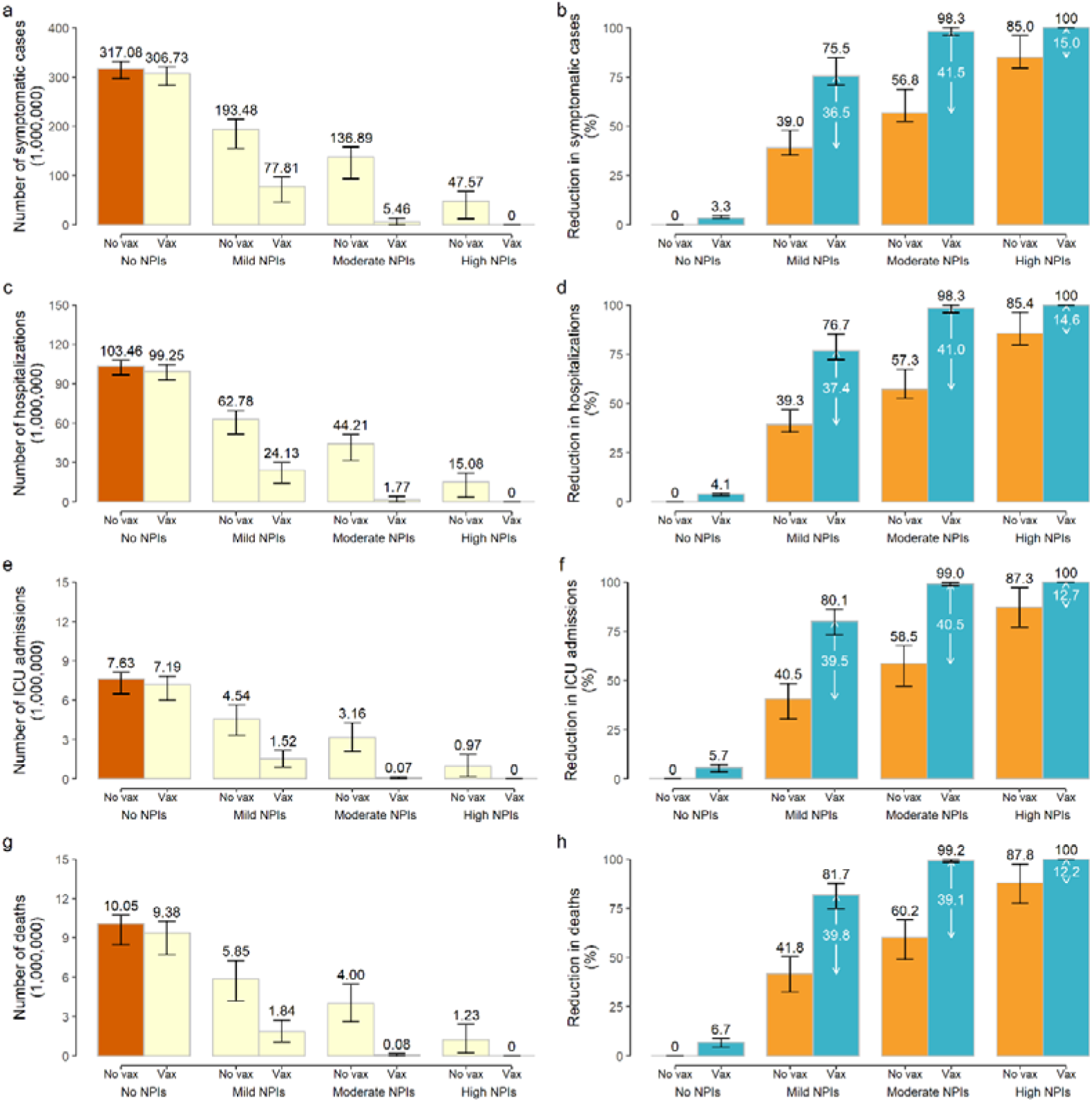
Burden of COVID-19 in the main analysis. a) Cumulative number of symptomatic cases as estimated under the different scenarios in the absence/presence of vaccination over the simulated 2-year period. No vaccination + no NPIs with R_t_=2.5 at the beginning of the outbreak is called *reference scenario*, described using dark brown bars. Light yellow bars indicate scenarios including vaccination and/or different levels of NPIs. b) Reduction in the cumulative number of symptomatic cases with respect to the *reference scenario*. Orange bars and black values indicate the contribution of NPIs, blue bars and black values indicate the overall contribution of vaccination and NPIs, while the white values indicate net contribution of vaccination; c)-d) As a-b but for hospitalized cases; e)-f) As a-b but for cases admitted to ICU; g)-h) As a-b but for deaths. Number denotes median, and error bars denote quantiles 0.025 and 0.975.

Provided that NPIs are in place and can keep R_t_ at 1.3 in the absence of vaccination (*moderate NPIs scenario*), the vaccination program could reduce COVID-19 burden by about 99% compared to the *reference no-vaccination scenario*, with 5.46 million (95%CI, 2.47-13.36) symptomatic cases, 1.77 million (95%CI, 0.83-4.40) hospitalizations, 73,500 (95%CI, 7,300-152,100) ICU admissions, and 76,700 (95%CI, 8,200-165,700) deaths (Fig. 2). In this context, vaccination decreases COVID-19 burden by about 40% (Fig. 2) compared to a situation with moderate NPI alone, and R_t_ falls below the epidemic threshold about 9 months after the epidemic start (Fig. 1). At the time that R_t_ falls below 1, we estimate that 50.8% of the total population would have been vaccinated, while 0.8% would have been naturally infected (Fig. 1g-i). This highlights that a relevant proportion of the population would be still susceptible to SARS-CoV-2 at that time. Although in the long-term vaccination can ultimately lead to the suppression of transmission, it is necessary to maintain NPIs for one year after the onset of vaccination. Indeed, if NPIs are relaxed from moderate (R_t_=1.3) to mild (R_t_=1.5) 9 months after vaccination start, the cumulative death toll could increase from 76,700 to 173,000. In contrast, a small increase in cumulative deaths from 76,700 to 81,700 is expected if this relaxation occurs one year after vaccination start, while earlier or more drastic relaxations of NPIs lead to substantial increases in deaths (Extended Data Fig.1-3).

A combination of more stringent NPIs (i.e., capable of keeping R_t_ =1.1) and vaccination (*vax + high NPIs* scenario) could suppress the epidemic, with <2,300 symptomatic cases, and <50 deaths on average. Although the majority of the reduction of COVID-19 burden is ascribable to NPIs in this case (over 85%), the deaths averted due to vaccination are about 1.2 million (Fig. 1j-l, and Fig. 2).

If we consider a set of mild NPIs (*vax + mild NPIs* scenario), even a relatively low initial reproduction number under NPIs of R_t_=1.5 could still lead to a disastrous epidemic, with nearly two million deaths. Despite the high death toll of the resulting epidemic, NPIs and vaccination would jointly reduce around 80% of the disease burden compared to a scenario with no NPIs and no vaccination (namely, 239 million symptomatic cases and 8.2 million deaths averted) (Fig. 1d-f, and Fig. 2).

### Vaccine distribution capacity

Should the daily vaccination rollout be limited to 1.3 million doses (1 per 1,000 individuals – a slower rate than during the 2009 H1N1 pandemic), vaccination would not effectively reduce COVID-19 related deaths unless there was adoption of stringent NPIs. In a scenario where vaccination capacity reaches 10 million doses administered per day (7 per 1,000 individuals), vaccination would reduce COVID-19 related deaths to <5,000 for moderate NPIs and <30 for high NPIs. Should the daily vaccination capacity be increased to 15 million doses (10 per 1,000 individuals), vaccination could effectively reduce deaths to <100,000 (similar to the annual influenza-related death toll in China^35^) even in the presence of mild NPIs. However, even if the daily vaccination capacity could be increased to 30 million doses (20 per 1,000 individuals), in the absence of NPIs, we estimate that over 7.7 million deaths would still occur (Fig. 3). Similar patterns are estimated for the number of symptomatic cases, hospitalizations and ICU admissions (Extended Data Fig.4-6).

**Figure 3.**
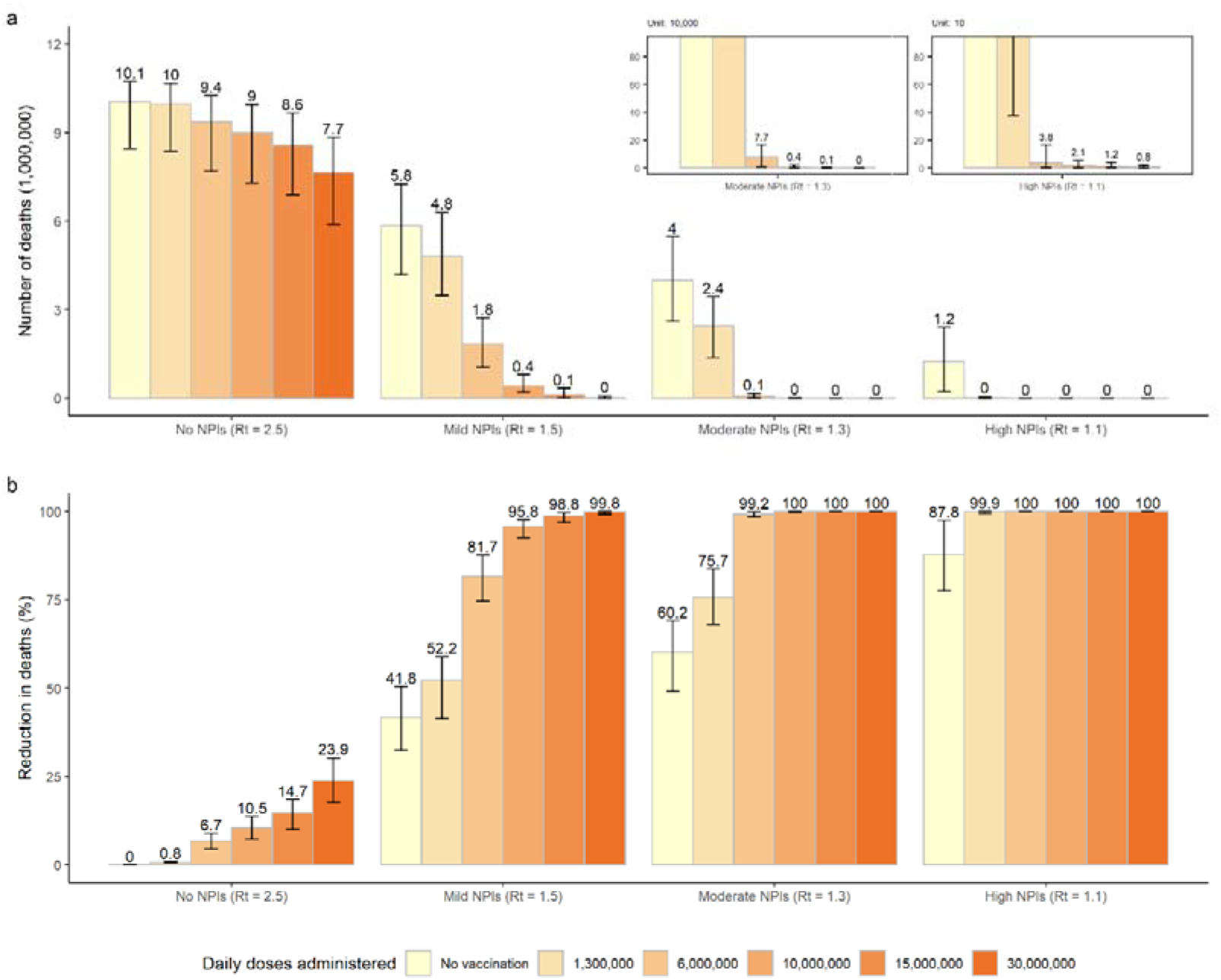
Impact of daily vaccine administration capacity on COVID-19 deaths. a) Cumulative number of COVID-19 deaths (millions) as estimated in the different scenarios under progressively increasing values of the daily vaccination capacity; b) Proportion of deaths averted compared to the *reference scenario*, i.e., *no vaccination + no NPIs* with R_t_=2.5 at the beginning of the outbreak. Number denotes median, and error bars denote quantiles 0.025 and 0.975.

**Figure 4.**
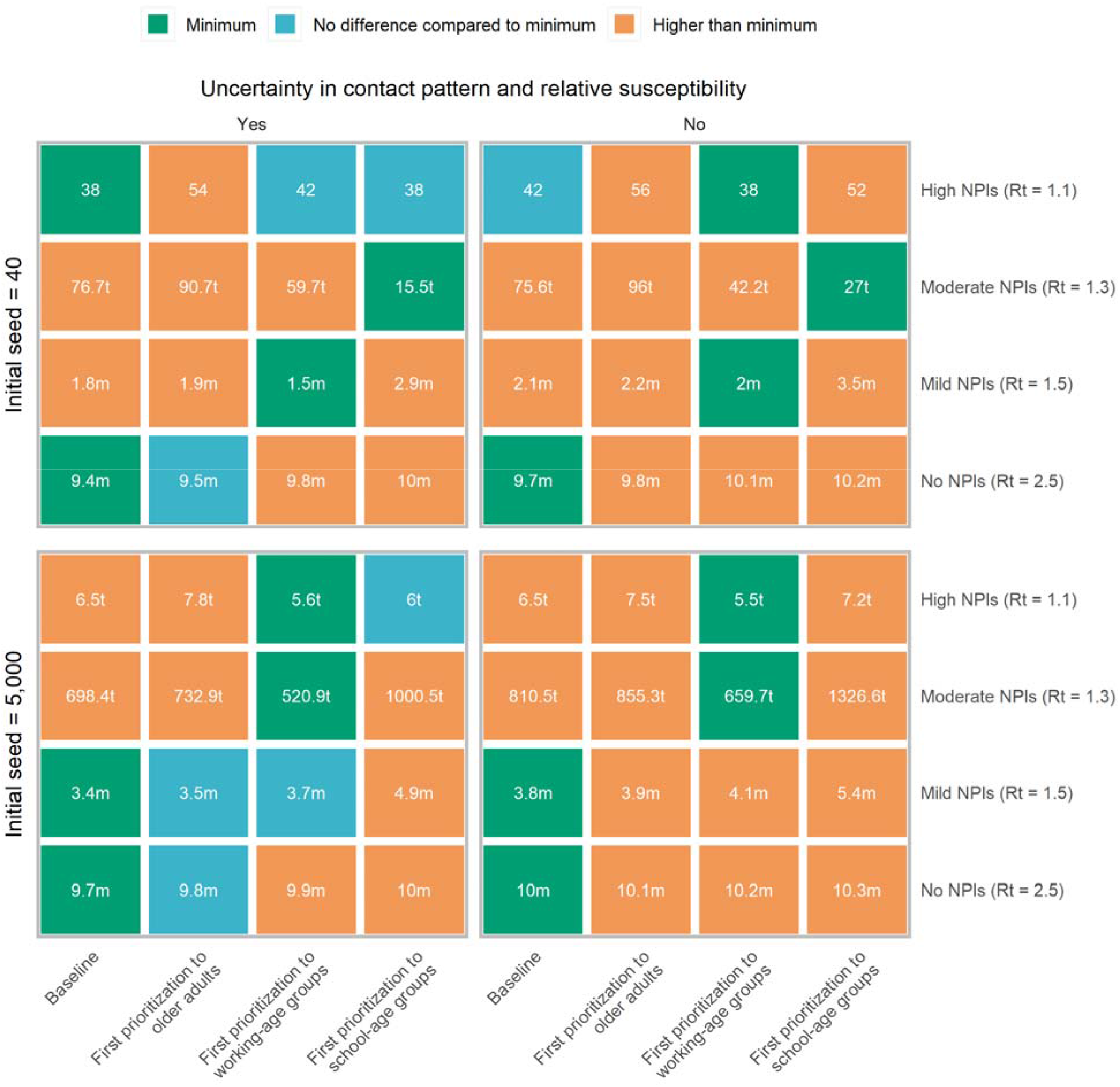
Best prioritization strategy to achieve the minimal COVID-19 deaths. Initial cases denote breakthrough COVID-19 cases, which initiates the epidemic. We consider the impact of uncertainty (i.e., “Yes” in the figure, which means 200 stochastic model realizations are performed in order to account for the ranges) in contact patterns and relative susceptibility on prioritization, and use their mean values as well (i.e., No). Baseline denotes first prioritizing older adults and individuals with underlying conditions. Number in the box denotes the death toll (median), with t representing thousand and m representing million. Minimum denotes the lowest deaths in each scenario on the basis of median value. We compare other strategies to that with minimum deaths using rank sum test. E.g., in the context of initial cases=5,000, R_t_=1.5 and using mean values of contact patterns and relative susceptibility, the baseline is the optimal strategy to minimize deaths.

Increasing daily vaccination capacity could largely shorten the time needed to control SARS-CoV-2 transmission. For instance, when considering a daily capacity of 10 million and 15 million doses and moderate NPIs, R_t_ would drop below 1 about 8 and 6 months respectively after epidemic onset (to be compared to the 9.3 months estimated with the baseline capacity of 6 million doses). At that time, over 60% of the population would be vaccinated and ≤0.1% would be naturally infected. An upscale in the daily capacity to 10 or 15 million doses would allow a relaxation of NPIs from moderate to mild already 6 to 9 months after vaccination start, i.e., 3 to 6 months earlier with respect to the baseline. On the other hand, more drastic relaxations of NPIs (e.g., from moderate to no NPIs) would still lead to substantial increases in symptomatic cases and deaths (Extended Data Fig.7-10).

### Vaccination prioritization

We consider alternative vaccination scenarios that prioritize essential workers (staff in the healthcare, law enforcement, security, community services, and individuals employed in cold chain, etc.) to maintain essential services and then explore different prioritization strategies for the rest of the population. Our results suggest that the relative timing of the epidemic and of the vaccination rollout play a key role in determining the most effective strategy. In particular if we consider vaccination to start two weeks after 40 cases are detected, there is no clear prioritization strategy that minimize deaths, as the outcome of the vaccination campaign heavily depends on the timing at which the epidemic unfolds (Fig. 4 and Extended Data Fig. 11-12). Instead, if the epidemic is already underway when the vaccination campaign starts (>5,000 cases), prioritizing working-age groups minimizes the number of deaths when R_t_≤1.3. In contrast, prioritizing older adults and individuals with underlying conditions is more effective when R_t_≥1.5 (direct benefits are higher, Fig. 4 and Extended Data Fig. 11-12). Two results are independent of the adopted prioritization strategy: i) if R_t_≥1.5, then an epidemic cannot be avoided; and ii) when R_t_ =1.1, over 99% of deaths can be averted (Extended Data Fig. 11-12).

### Population immunity at the onset of an outbreak

In December 2020, vaccination has started in China, while measures to detect imported cases and case surveillance are in place. The number of doses distributed per day has changed over time, following an increasing trend and with a daily average of about 6 million doses over the period April 20 – May 7, 2017 (Supplementary File 3). As of May 7, 2021, the vaccination coverage is 10.7% ^6^. The effectiveness of the vaccination program and NPIs in preventing new COVID-19 outbreaks and limiting COVID-19 burden will thus depend on the level of vaccine-induced immunity in the population should an outbreak of locally transmitted cases start to unfold.

To simulate this situation, we initialize the model by considering different fractions of vaccinated population (*SA1*: 10%; *SA2*: 20%; *SA3*: 30%) at the time the infection is seeded. Given a certain level of NPIs in the absence of immunity, increasing proportions of vaccinated individuals will decrease the effective reproduction number (e.g., R_t_ =1.1 in the absence of immunity, corresponds to R_t_ below the epidemic threshold if 10% or more of the population has been vaccinated).

Should 30% of the population already been vaccinated before the start of a new wave, continuing the vaccination program while adopting mild NPIs would reduce the death toll by 98% (42,400 deaths as compared 1.8 million if no one was vaccinated). However, in the absence of NPIs, even if 30% of population was already vaccinated before the start of a new wave, carrying on with the vaccination program alone would not be enough to prevent a widespread epidemic leading to 6 (95%CI: 4-7) million deaths (Extended Data Fig.13).

### Vaccination coverage

No significant difference in willingness-to-vaccination between age groups has been reported in China ^36–38^. Accordingly, we use a homogenous vaccine coverage of 70% among all age groups in the main analysis. Here we present the results of a set of sensitivity analyses assuming vaccination coverage of 50% (*SA4*) and 90% (*SA5*) among all age groups, and considering heterogeneous coverages by age: 1) 70% for adults ≥20 years and 50% for younger individuals (*SA6*); 2) 90% for adults ≥20 years and 70% for younger individuals (*SA7*); 3) 70% for adults ≥20 years and no vaccination for younger individuals (*SA8*).

By considering moderate NPIs (R_t_=1.3) and a vaccination coverage of 50% in all age groups, the number of symptomatic is estimated to decrease by 33% with an 8% decrease of the death toll with respect to the baseline vaccination scenario (70% coverage) (Extended Data Fig.14-15). In fact, the lower coverage in older age groups would lead to an earlier vaccination in younger age groups who are characterized by the highest contact rates ^13^. Conversely, increasing the coverage to 90% would decrease the death tolls but lead to an increase of symptomatic cases (Extended Data Fig.14-15). It is important to remark that these results consider that moderate NPIs remain in place over the entire duration of the epidemic, and they do not provide indications of the coverage needed to suppress any further resurgences of cases.

As compared to the baseline vaccination scenario (70% vaccination coverage in all age groups), if the vaccine is not distributed among individuals aged less than 20 years, we estimate an increase of the death toll of 69% and of symptomatic infections of 55% (Extended Data Fig.14-15). The scenario assuming 90% vaccination coverage for adults ≥20 years and 70% for younger individuals lead to a lower reduction of COVID-19 burden as compared with the baseline vaccination scenario (Extended Data Fig. 14-15). The higher coverage in the adult population result in a delayed start of vaccination of the young population, which is characterized by higher contacts rates ^13^. Nonetheless, it is important to remark that this result depends on the timing of the introduction of the initial seeds (see previous section).

### Vaccine efficacy

Three types of COVID-19 vaccines are currently in use in China, including inactivated, recombinant protein subunit, and adenovirus-vectored vaccines. VE for these vaccines ranges from 65% to 90%, with the exception of the one tested in Brazil where P1 variant is prevalent (VE=50%) ^39–43^. With respect to an 80% VE adopted in the baseline scenario, by considering VE=60% (*SA9*) we estimate a 1.02-fold increase of the death toll and 1.64-fold increase of symptomatic cases. Whereas a 37% and 29% decrease of deaths and symptomatic cases, respectively, is estimated for VE=90% (*SA10*). (Extended Data Fig.14-15)

### SARS-CoV-2 variants

Multiple SARS-CoV-2 variants have been documented globally, three of which are of particular concern: lineage B.1.1.7 identified in UK, B.1.351 in South Africa, and P.1 in Brazil ^44^. These variants are estimated to have higher transmissibility ^45, 46^ and possibly an increased mortality ^47, 48^. To assess the effect of vaccination in this context, we consider higher values of R_t_ (i.e., 1.7, 1.9, and 2.1 – about 30%-60% increased transmissibility with respect to the main analysis ^45^) to account for enhanced transmissibility, and use a mean death hazard ratio of 1.64 to account for higher mortality ^47^. With the assumption of VE=80% against the new variant, COVID-19 burden substantially increases, compared to the scenario based on the historical lineage. The number of symptomatic cases increase from 2 thousand to 173 million, and death increase from <50 to 7 million even when strict NPIs are implemented (Extended Data Fig. 16).

### Alternative vaccination parameters and scenarios

A further set of sensitivity analyses are conducted to evaluate the impact of baseline assumptions on our results for R_t_=1.3 (moderate NPIs). Provided that vaccination can only protect against illness (*SA11*) but not SARS-CoV-2 infections, COVID-19 related deaths increase by 33 folds with respect to the baseline: from 76,700 to 2.66 million (Extended Data Fig.14). In this case, maintaining stringent NPIs measures in place for a prolonged time horizon would be necessary as such vaccine would not be effective to suppress transmission (as reported in previous studies ^49^). Assuming a shorter duration of vaccine-induced protection of 6 months (*SA12*) instead of a lifelong protection (i.e., longer than the 2-year time horizon considered, Extended Data Fig.14) has a similarly large effect on projections.

In our main analysis, we use the contact matrix estimated from a contact survey conducted in Shanghai before the COVID-19 pandemic ^13^. Should a new COVID-19 wave start to unfold in China, it is unclear to what extent pre-pandemic contact patterns could be representative of such a situation. Therefore, we have added a sensitivity analysis where we assume the mixing patterns estimated in Shanghai in March 2020 ^50^, when schools were still closed as a response to the COVID-19 pandemic (*SA13*). For R_t_=1.3 and the baseline parameters for the vaccination, the estimated number of deaths would be 16,000 as compared to 76,700 estimated using the pre-pandemic mixing patterns (79% decrease, see Extended Data Fig.14). In fact, the relative contribution of the adult population (which is the main target of the vaccination campaign) to the overall transmission as compared to children is higher than when considering pre-pandemic mixing patterns (when schools were open and school-age individuals had the highest number of contacts).

Other factors such as excluding detected symptomatic cases from vaccination (*SA14* and *SA15*), the time interval between two doses (*SA16 and SA17*), and assuming an all-or-nothing vaccine (*SA18*), do not substantially affect estimates of deaths and symptomatic infections (Extended Data Fig.14-15). A similar trend is observed for hospitalized cases and ICU admissions.

## Discussion

Using a stochastic dynamic model of SARS-CoV-2 transmission and COVID-19 burden tailored to the epidemiological situation in China, we find that in the absence of NPIs, and independently of the vaccine prioritization strategy and capacity of the vaccination campaign, timely rollout of an effective vaccine (VE =80%) would not be enough to prevent a local outbreak to escalate to a major widespread epidemic.

Provided that NPIs are in place and capable to bring R_t_ to 1.3, a daily vaccine rollout of 4 doses per 1,000 individuals could reduce around 99% of COVID-19 burden, and bring R_t_ below the epidemic threshold about 9 months after the start of the vaccination campaign. A relaxation of NPIs that bring the value of R_t_ to 1.5 could not prevent sustained epidemic growth which would cause 1.8 million deaths. A net reproduction number of 1.5 could only be sustained when accompanied by an improvement of the vaccine administration capacity up to 10 doses per 1,000 individuals per day. Relaxation of NPIs in the first 6-9 months of vaccine rollout could lead to substantial increases of COVID-19 burden if daily vaccination capacity could not be enhanced to 10-15 million doses.

Bubar K, et al. evaluated COVID-19 vaccine prioritization strategies and found that prioritizing older adults is a robust strategy to minimize deaths across countries when R_t_=1.5, while prioritization shifted to 20-49 years group when R_t_=1.15^51^. The broad scope of that multi-country analysis does not account for features of COVID-19 epidemiology and vaccination program that are unique to China. In particular, differently from most countries where natural immunity is building up after widespread epidemics, China has been able to suppress SARS-CoV-2 transmission for most of 2020. As a result, prior immunity is very low, thus calling for specifically tailored analysis. Nonetheless, our findings confirm that if NPIs can maintain transmission rates at low levels during the vaccination campaign, strategies that target indirect benefits perform better, while if transmission rates remain high, strategies maximizing direct benefits may save more lives^51^.

As highlighted in vaccination studies in Italy^52^, in the race between the vaccination campaign to build population herd-immunity and the progress of the epidemic, the speed of vaccine deployment is critical. Considering the average vaccine distribution capacity of the current COVID-19 vaccination campaign in China ^6^, we use 6 million doses administered per day in the baseline analysis. Several manufacturers state that a total of 3.9 billion doses of COVID-19 vaccine could be produced in 2021, equivalent to about 10 million doses per day ^22–26^. China committed to provide COVID-19 vaccines to >100 countries, and thus that could reduce the number of doses to be distributed locally. Even if these candidate vaccines could be licensed and manufactured smoothly, it would take about one year to vaccinate 70% of the population.

Five months after initiating vaccination program, 10.7% of Chinese population has been vaccinated ^6^. Limited vaccine production capacity, particularly at the initial stage, could slow the speed of vaccine rollout. Slower rates of vaccine production and administration may result in a longer period of SARS-CoV-2 transmission. It is thus crucial to keep monitoring local outbreaks and invest resources in outbreak management (as currently done in China) in order to keep R_t_ close to the epidemic threshold at least for the next 1-2 years. In the very unique context of China, a value of R_t_ of 1.3 would result in about 76,700 cumulative deaths, comparable to the annual influenza-related death toll in China^35^. The development of detailed logistical plans and tools to support an increased vaccination capacity as well as effective logistic (vaccine transport, storage, and continuous cold-chain monitoring) are key factors for a successful mass vaccination campaign.

In the early phase of COVID-19 spread in Wuhan in 2019, before interventions were put in place, R_0_ was estimated to be in the range 2.0-3.5^27–32^. Given the knowledge of mechanisms of SARS-CoV-2 transmission, and the devastating consequences of an uncontrolled COVID-19 epidemic, the Chinese population would hold a precautious behavior (such as clean hands often, cough or sneeze in bent elbow, avoid close contact with someone who is sick, etc.) even without the need to impose NPIs. As such, in our analysis simulating an epidemic triggered by imported cases, we decided to consider an initial R of 2.5, which is at the lower end of the estimated spectrum.

SARS-CoV-2 variants are circulating globally and quickly became dominant in countries like the UK and Italy (lineage B.1.1.7), and South Africa (lineage B.1.351). Recently, a variant B.1.617 identified in India has raised global concern. As of May 7, 2021, Mainland China border control screenings have already identified imported cases with SARS-CoV-2 lineage B.1.1.7 and B.1.617. Our study shows that the spread of new more transmissible and/or more lethal variants could substantially decrease the net benefit of vaccination. Strict border quarantine and isolation as well as genomic surveillance will be key while vaccines roll out in China.

Our analysis on the vaccine efficacy shows that if we consider VE=60%, both the number of symptomatic cases and deaths are estimated to double as compared to the baseline vaccination coverage of 80%. Given that the final composition of a nation-wide rollout will likely include a combination of vaccines with varying efficacy, monitoring vaccine effectiveness on the ground will remain a priority.

Here we propose a general framework to evaluate the impact of COVID-19 vaccination programs in the absence/presence of NPIs and to explore priority target populations to minimize multiple disease outcomes. The proposed modeling framework is adaptable to other country-specific contexts. However, it requires the collection of country-specific data about the epidemiological situation (e.g., landscape immunity of the local population, prevalence of infections), vaccination parameters (e.g., vaccine supply and capacity of immunization services, efficacy of different vaccines, target age groups), socio-demographic characteristics of the population (e.g., size of the priority population by age group, age-mixing patterns), and the priorities of the pandemic responses (e.g., limit the death toll, prevent infections).

Our study has a number of limitations. First, we integrated the impact of NPIs through a simple reduction in the value of R_t_ at the beginning of the outbreak, homogeneously across age groups. However, our analysis does not suggest which combination of NPIs should be adopted to lower R_t_ to a certain level, and how this would affect transmission rates in different age groups. Li, et al, estimated that individual NPIs, including school closure, workplace closure, and public events bans, were associated with reductions in R_t_ of 13–24% on day 28 after their introduction ^53^. Further studies are needed to pinpoint the specific NPIs to be adopted in parallel with the vaccination campaign and their impact on the quality of life of the population.

Second, in China, vaccines have not been licensed for children, so we assume a 50% lower or equivalent VE for them compared to other adults. Although we show that variations in these rates do not substantially affect the overall effect of the vaccination campaign, further data on age-specific vaccine efficacy could help refine priority groups. Our sensitivity analyses on vaccine coverage reveal the importance of extending the vaccination to the young population once the use of vaccines will be authorized for that age segment of the population.

Third, we assumed that immunity after natural infections lasts more than the time horizon considered (two years). If this is not the case, waning of immunity would inflate the rate of susceptible individuals and thus require booster vaccinations. This could become an issue with the emergence of immune-escape variants, as reported in South Africa^54^. Given limited information at this stage, we did not consider this scenario in our analyses, but this is an important area of future research.

Fourth, age-mixing patterns are key to assess the impact of vaccination as individuals of different ages are exposed to different transmission risks. In the main analysis, we assumed the mixing patterns to correspond to those estimated before the COVID-19 pandemic, indicating the goal of a return to pre-pandemic interactions. We have also performed a sensitivity analysis based on the mixing patterns estimated in China in March 2020^50^, after the lockdown was lifted but schools were still closed. How the population would mix in case of a new wave of COVID-19 starts to unfold in China remains to be seen.

Moreover, our study is performed at a national scale and thus our estimates of the impact of vaccination should be interpreted cautiously at the local scale. In fact, spatial heterogeneities within China in terms of risk of case importation, socio-demographic characteristics of the population, mixing and mobility patterns, vaccination coverage and capacity may affect our results.

Enhanced vaccination efforts in conjunction with NPIs have been successfully used during the COVID-19 outbreak in Ruili city (Yunnan Province, China) in March-April, 2021. Our analysis, however, focuses on the assessment of whether and to what extent we need to rely on NPIs to prevent a COVID-19 epidemic while vaccines are rolled out. As such, our results cannot be used to guide a reactive spatially targeted strategy. To properly capture the peculiarity of that context, specific modeling tools mirroring the interventions adopted in China as a response to emerging outbreaks are needed.

Finally, it would be interesting to analyze adaptive vaccination prioritizations that changes as the epidemiological situation evolves over time but that would require the development of dynamic optimization algorithms that are beyond the scope of this work ^55^. Nonetheless, our study provides estimates of the effect of relaxing NPIs over the course of the epidemic.

In conclusion, vaccination alone could substantially reduce COVID-19 burden, but in the foreseeable future may not be enough to prevent local outbreaks to escalate to major widespread epidemics due to limitation in the vaccine production and supply (particularly at the initial stage of the vaccination), as well as the capacity of vaccination system. This is especially relevant in contexts where most of the population is still susceptible to SARS-CoV-2 infection, as it is the case in most of China. Maintaining NPIs (such as social distancing, testing, case isolation and contact tracing, wearing masks, and limitation on large gatherings) throughout 2021 is necessary to prevent the resurgence of COVID-19 epidemics until a sufficiently high level of immunity is reached, which depends on the transmissibility of the variants circulating at that time.

## Data Availability

Should the manuscript be accepted, all data and codes will be provided on GitHub.

## Methods

### SARS-CoV-2 transmission and vaccination models

We developed a model of SARS-CoV-2 transmission and vaccination, based on an age-structured stochastic susceptible-infectious-removed (SIR) scheme, accounting for heterogeneous mixing patterns by age as estimated in Shanghai ^13^. The Chinese population was distributed in 18 age groups (17 5-year age groups from 0 to 84 years and one age group for individuals aged 85 years or older) ^57^. Each age group was further split into two subgroups: individuals with or without underlying conditions, where the former was considered to be associated with an increased risk of severe outcome of COVID-19 ^14^.

In the main analysis, susceptibility to SARS-CoV-2 infection was assumed to be heterogeneous across ages. Children under 15 years of age were considered less susceptible to infection compared to adults aged 15 to 65 years, while the older adults more susceptible ^33, 34^. Homogeneous susceptibility across age groups was explored in sensitivity analysis (***SA19***). Asymptomatic and symptomatic individuals were assumed to be equally infectious ^33, 34^, and infectiousness was also assumed to be the same across age groups ^33, 34^.

Vaccine is administered with a two-dose schedule. In the baseline model, we assumed that: i) vaccination reduces susceptibility to SARS-CoV-2 infection; ii) only susceptible individuals are eligible for vaccination, i.e., we excluded all individuals that have experienced SARS-CoV-2 infection; iii) duration of vaccine-induced protection lasts longer than the time horizon considered (2 years).

The baseline model is schematically represented in Supplementary Fig.1 and it is described by differential systems presented in Supplementary File 1-2.

### Model initialization

In China, the first pandemic wave of COVID-19 was controlled by intense NPIs ^58, 59^. Almost the entire population of mainland China is still susceptible to COVID-19 ^3^. As such, the model is initialized with a fully susceptible population.

China has been facing mounting pressure of imported COVID-19 cases. Containment of COVID-19 has been possible only through a combination of measures such as complete- or partial-lockdown, citywide mass-screening using reverse-transcriptase–polymerase-chain-reaction (RT-PCR) testing, tracing of contacts and contacts of contacts of COVID-19 cases, which were promptly applied wherever COVID-19 transmission has popped up in mainland China ^60^. Despite all the efforts, containment of COVID-19 appears a whack-a-mole game and sporadic outbreaks inevitably occur. Simulations are thus initialized with 40 cases, roughly corresponding to the number of cases with symptoms onset in Beijing before the detection of a local outbreak in June 11, 2020 ^61^.

### Vaccination scenarios

To explore the impact of vaccination, we ran a set of simulations in which neither NPIs nor vaccination are implemented as a *reference scenario* (*no vax + no NPIs*, i.e., effective reproductive number R_t_=2.5 at the beginning of simulations ^17, 28, 58^), and compared it with a scenario in which vaccination only is implemented (*vax + no NPIs*). Further, we considered different sets of simulations in which NPIs are used to bring R_t_ respectively down to 1.5 (mild NPIs), 1.3 (moderate NPIs), and 1.1 (high NPIs), with (*vax + mild/moderate/high NPIs*) or without vaccination program (*no vax* + *mild/moderate/high NPIs*). In the main analysis vaccination is assumed to begin 15 days after the epidemic start. Alternative scenarios about the seeding of the epidemic were explored as sensitivity analyses. In particular, we considered the epidemic to start when 10% (***SA1***), 20% (***SA2***), and 30% (***SA3***) of the Chinese population has already been vaccinated.

The model is run considering daily time steps. Gradual delivery of vaccine doses is implemented by vaccinating a fixed number of individuals each day. Although manufacturers state that a total of 3.9 billion doses of vaccines could be available by the end of 2021 ^22–26^, scale-up and delivery will take months. On the basis of the 2009 H1N1 influenza pandemic vaccination program implemented in mainland China ^62^, in the main analysis we assumed 6 million doses of COVID-19 vaccines could be administered each day (4 doses per 1,000 individuals) until uptake reaches 70% for all groups ^14^. Different values of the daily vaccine administration capacity, i.e., 1.3 (***SA20***), 10 (***SA21***), 15 (***SA22***), 30 (***SA23***) million dose per day, are explored in separate sensitivity analyses. Sensitivity analyses were also performed on the vaccination coverage which is assumed either to be homogenous (***SA4*** and ***SA5***)^14^ or heterogeneous by age (***SA6***, ***SA7***, and ***SA8***).

In the main analysis, vaccination is administered to susceptible individuals only. This represents an ideal scenario where we assume that all infected individuals can be identified (e.g., either via RT-PCR while infected or via serological assays later one) and that SARS-CoV-2 infection confers a long-lasting immunity. Since infection ascertainment could be challenging and pose additional strain to the health system, we also consider two sensitivity analyses in which only detected symptomatic cases are excluded from vaccination (***SA14-SA15***).

In the context of fast RT-PCR–based mass screening if there is an outbreak, under-ascertainment of symptomatic cases could be only related with the sensitivity of RT-PCR tests. The sensitivity is quite high (98%) if the interval between symptom onset and RT-PCR test is within 7 days, and the sensitivity decreases to 68% if the time interval is 8-14 days^63^. The mean time interval from symptom onset to the date of collection of the sample for PCR testing was estimated to be 4.7 days in Hunan^33^. Accordingly, we considered as ascertainment probabilities of symptomatic cases 70% (***SA14***) and 90% (***SA15***).

### Vaccination schedule and efficacy

There are six COVID-19 vaccines developed by China in phase 3 clinical trials, including five vaccines administered with a two-dose schedule with an interval of 14, 21, or 28 days and one single-dose recombinant adenovirus type-5-vectored vaccine. For simplicity, in the main analysis, we modeled the administration of an inactivated vaccine developed by the Beijing Institute of Biological Products,^64^ which entail a two-dose schedule across all age groups with an interval of 21 days. In separate sensitivity analyses, we explored an interval of 14 days and 28 days (***SA16****-**SA17***).

China approved its first local COVID-19 vaccine (developed by Sinopharm) for general public use on December 31, 2020, with an estimated vaccine efficacy (VE) of 79.3%.^21^ In the main analysis, we used a VE of 80% against infection in individuals aged 20-59 years. In the developed model, vaccination confers a partial protection, i.e., vaccinated individuals are 80% less likely to develop infection upon an infectious contact. Sensitivity analyses using a VE of 60% (***SA9***) and 90% (***SA10***) were separately performed. The alternative values of VE were selected on the basis of published upper efficacy of vaccines of 94-95% and in such a way to cover a plausible efficacy range of forthcoming vaccines ^65–67^.

Phase 2 clinical trials demonstrated that vaccine immunogenicity was lower among older individuals than in younger adults ^64^. And for other inactivated vaccines like influenza vaccine, a lower VE is observed in children compared to young adults ^68^. Accordingly, we assumed an age-dependent VE. In particular, given a baseline efficacy VE among individuals aged 20-59 years (80% in the main analysis), we assumed a 50% lower VE in individuals <20 and ≥60 years of age (namely 40%). A scenario without age-specific variations in VE was explored as sensitivity analysis (***SA24***).

Individuals vaccinated with the first dose could still develop infections without any immune protection, while the second dose vaccination could produce the expected vaccine efficacy after an average of 14 days. In the main analysis we assume both natural infection-induced and vaccine-induced immunity to SARS-CoV-2 infection does not wane within the considered time horizon (2 years). In additional sensitivity analyses, we considered an average duration of vaccine-induced protection of 6 months (***SA12***) and 1 year (***SA25***). We also consider a sensitivity analysis assuming that vaccination is effective in preventing symptomatic illness, but not infection (***SA11***), and another one assuming an all-or-nothing vaccine, i.e., the vaccine confers full protection to VE percent of vaccinated individuals (***SA18***).

### Priority order of vaccination

The doses available to be distributed daily (6 million in the main analysis) are assigned by considering the following order of priority ^14^. In the main analysis, healthcare workers are considered as the top priority (Tier 1 of the vaccination strategy); law enforcement and security workers, personnel in nursing home and social welfare institutes, community workers, workers in energy, food and transportation sectors are included in Tier 2; adults ≥60 years of age with underlying conditions, and adults ≥80 years of age without underlying conditions, who are at the highest risk of severe/fatal COVID-19, are considered in Tier 3; individuals aged < 60 years with pre-existing medical conditions and pregnant women are included in Tier 4; individuals aged 20-59 years without underlying conditions are included in Tier 5; school-age children and younger children aged ≤5 years without underlying conditions are recommended for vaccination in Tier 6 (Supplementary File 4).

Different priority orders are explored as sensitivity analyses. Healthcare workers and the other essential workers listed above are fixed in Tier 1 and Tier 2 of vaccination, while the remaining population is vaccinated as described in Supplementary Table 2 by considering different orders of prioritization only based on age and disregarding the presence of underlying conditions (***SA26***: first prioritization to old adults; ***SA27***: first prioritization to working-age groups***; SA28***: first prioritization to school-age groups). We explore the impact of 5,000 initial cases on the prioritization strategy (***SA29***). To understand the impact in terms of number of infections by age, we compare the prioritization strategy when we account for the uncertainty in the contact matrix and in the susceptibility to infection by age, or not (in this context, median values of contact numbers and relative susceptibility are used).

### COVID-19 burden model

The main output of above transmission model is the age-specific number of new infections per day in the subpopulation with or without underlying conditions. On top of that, we developed a model of COVID-19 disease burden to estimate the number of symptomatic cases, hospitalization, ICU admissions, and deaths in different scenarios in the presence/absence of vaccination.

We computed the age-specific number of symptomatic infections in individuals with and without underlying conditions on a daily-basis, by applying an age-specific probability of respiratory symptoms, which is 18.1%, 22.4%, 30.5%, 35.5%, and 64.6% separately for 0-19, 20-39, 40-59, 60-79, and 80+ years of age, as estimated from contact tracing data in Lombardy ^20^. We assume that individuals with and without underlying conditions have the same age-specific probability of developing symptoms.

The daily age-specific number of hospital admissions in the two subpopulations was computed by applying the age-specific proportion of laboratory-confirmed symptomatic cases requiring hospitalizations (Supplementary File 5), delayed by an average time of 3.8 days between symptom onset and hospitalization ^17^.

The daily age-specific number of patients admitted to ICU in the two subpopulations was computed by applying to hospitalized cases an age-specific probability of being admitted to ICU ^19^, and distinguishing patients requiring intensive care in survivors and non-survivors. Survivors are admitted to ICU after an average time of 7 days from hospitalization. Non-survivors are admitted to ICU after an average time of 8 days after hospitalization ^16^.

The daily age-specific number of deaths in the two subpopulations was computed by applying the age-specific fatality ratio among symptomatic cases (Supplementary File 5), delayed by an average time of 13.9 days between symptom onset and death ^18^.

### Data analysis

For each scenario, 200 stochastic model realizations were performed. The outcome of these simulations determined the distributions of the number of symptomatic infections, hospitalizations, ICU admissions, and deaths. 95% confidence intervals were defined as quantiles 0.025 and 0.975 of the estimated distributions. We used a Bayesian approach to estimate R_t_ from the time series of symptomatic cases by date of symptom onset and the distribution of the serial interval ^17^. The methods were described in Supplementary File 6 in details.

### Ethics approval

All these data were in the public domain. Ethical review for the re-use of these secondary data is not required.

### Reporting summary

Further information on research design is available in the Nature Research Reporting Summary linked to this paper.

### Data and code availability

Should the manuscript be accepted, all data and codes will be provided on *GitHub*.

## Acknowledgments

The study was supported by grants from the National Science Fund for Distinguished Young Scholars (No. 81525023), Key Emergency Project of Shanghai Science and Technology Committee (No 20411950100), European Union Grant 874850 MOOD (MOOD 000).

## Author Contributions

H.Y. conceived the study. H.Y., S.M., and M.A. designed and supervised the study. J.Y., J. Z., J.C., W.W., Q.W., W.Z., Z.Z, K.D., and G.Z. participated in data collection. V.M., G.G., P.P., and F.T. developed the model. V.M., J.Y., and X.D. analyzed the model outputs and prepared the tables and figures. J.Y. prepared the first draft of the manuscript. H.Y., V.M., and M.A. commented on the data and its interpretation, revised the content critically. All authors contributed to review and revision and approved the final manuscript as submitted and agree to be accountable for all aspects of the work.

## Disclaimer

This article does not necessarily represent the views of the NIH or the US government.

## Additional information

**Supplementary Information** is available for this paper.

**Correspondence and requests for materials** should be addressed to M.A., and H.Y.

## Peer review information

**Reprints and permissions information** is available at http://www.nature.com/reprints.

