## Supplementary Information for "To what extent do we need to rely on non-pharmaceutical interventions while COVID-19 vaccines roll out in 2021?"

### Supplementary file 1. SARS-CoV-2 transmission and vaccination models

We developed a model of SARS-CoV-2 transmission and vaccination, based on an age-structured stochastic susceptible-infectious-removed (SIR) scheme, accounting for heterogeneous mixing patterns by age as estimated in Shanghai ^1^. The Chinese population was distributed in 18 age groups (17 5-year age groups from 0 to 84 years and one age group for individuals aged 85 years or older) ^2^. Each age group was further split into two subgroups: individuals with or without underlying conditions, where the former was considered to be associated with an increased risk of severe outcome of COVID-19 ^3^.

In the main analysis, susceptibility to SARS-CoV-2 infection was assumed to be heterogeneous across ages. Children under 15 years of age were considered less susceptible to infection compared to adults aged 15 to 65 years, while the elderly more susceptible ^4^. Asymptomatic and symptomatic individuals were assumed to be equally infectious ^4,5^, and infectiousness was also assumed to be the same across age groups ^4,5^.

Vaccine is administered with a two-dose schedule. In the baseline model, we assumed that: i) vaccination reduces susceptibility to SARS-CoV-2 infection; ii) only susceptible individuals are eligible for vaccination, i.e., we excluded all individuals that have experienced SARS-CoV-2 infection; iii) duration of vaccine-induced protection lasts longer than the time horizon considered (2 years).

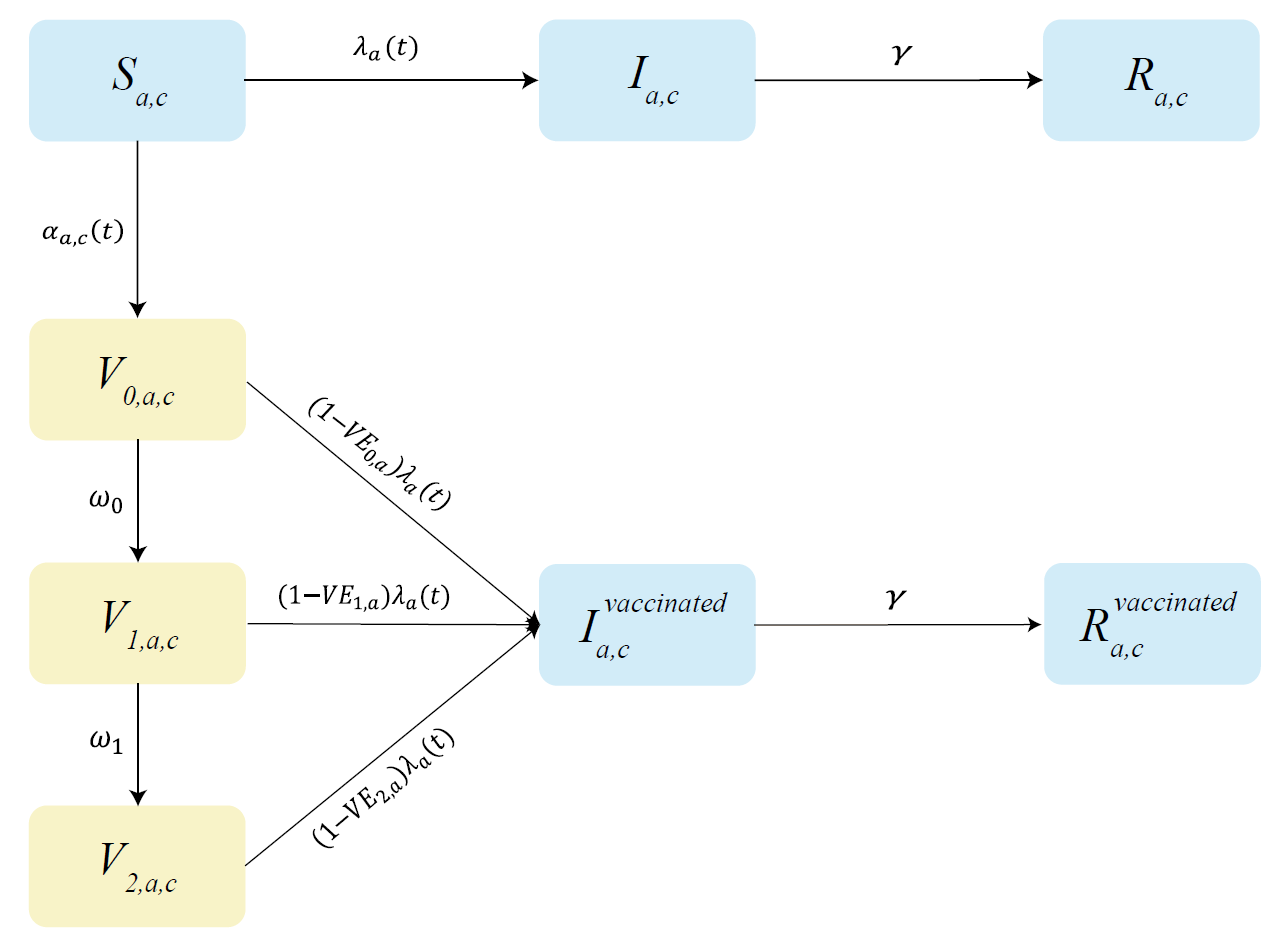

**Fig. 1 Schematic representation of the baseline model.**

Blue, and yellow rectangles describe the SARS-CoV-2 transmission model, and vaccination model, respectively. Transitions occur within each population class defined by a and c, where a represents the age group and c identifies the absence/presence of underlying conditions, the latter associated with higher risk of sever outcomes of SARS-CoV-2 infections. S denotes susceptible individuals; I infected individuals; R recovered/removed individuals. Parameters of the transmission model include$:$the time- and age-dependent force of infection$\lambda_{a}\left( t \right)$ and the recovery rate from infection ($\gamma)$. V_0_ denotes individuals vaccinated with the 1^st^ dose (never experienced infection with SARS-CoV-2); V_1_ denotes vaccinated with the 2^nd^ dose (no protection yet); V_2_ denotes vaccinated with the 2^nd^ dose (protected). Parameters of the vaccination model include: the time-, age- and group-dependent probability of being vaccinated $(\alpha_{a,c}\left( t \right))$; the interval between administration of the first and second dose (1/$\omega_{0}$); the delay of ramp-up of the 2^nd^ dose (${1/\omega}_{1}$); the age-dependent vaccine efficacy after the 1^st^ dose of vaccination (${VE}_{0,a})$, which is assumed to be 0; the age-dependent efficacy right after administration of the 2^nd^ dose (${VE}_{1,a}),$which is assumed to be 0; the vaccine efficacy after ramp-up of the 2^nd^ dose (${VE}_{2,a})$.

The baseline model is schematically represented in Fig. S1 and it is described by the following differential systems:

$$\left\{ \begin{aligned} S_{a,c}^{'}\text{(t)= }{-\lambda}_{a}\left( t \right)S_{a,c}\left( t \right)-\alpha_{a,c}\left( t \right)S_{a,c}\left( t \right) \\ I_{a,c}^{'}\text{(t)= }\lambda_{a}\left( t \right)S_{a,c}\left( t \right)-\gamma I_{a,c}\left( t \right) \\ R_{a,c}^{'}\text{(t)= }\gamma I_{a,c}\left( t \right) \\ V_{0,a,c}^{'}\text{(t)=}\alpha_{a,c}\left( t \right)S_{a,c}\left( t \right)-\left( 1{-VE}_{0,a} \right)\lambda_{a}\left( t \right)V_{0,a,c}-{\omega_{0}V}_{0,a,c}\left( t \right) \\ V_{1,a,c}^{'}\text{(t)=}{\omega_{0}V}_{0,a,c}\left( t \right)-\left( 1{-VE}_{1,a} \right)\lambda_{a}\left( t \right)V_{1,a,c}-{\omega_{1}V}_{1,a,c}\left( t \right) \\ V_{2,a,c}^{'}\text{(t)=}{\omega_{1}V}_{1,a,c}\left( t \right)-\left( 1{-VE}_{2,a} \right)\lambda_{a}\left( t \right)V_{2,a,c} \\ I_{a,c}^{\mathrm{vaccinated} '}\text{(t)=}\lambda_{a}\left( t \right)[\left( 1{-VE}_{0,a} \right)V_{0,a,c}+\left( 1{-VE}_{1,a} \right)V_{1,a,c}+\left( 1{-VE}_{2,a} \right)V_{2,a,c}]-\gamma I_{a,c}^{\mathrm{vaccinated}}\text{(}\text{t}\text{)} \\ R_{a,c}^{\mathrm{vaccinated} '}\text{(t)=}\gamma I_{a,c}^{\mathrm{vaccinated}}\text{(}\text{t}\text{)} \end{aligned} \right.$$

where:

$S_{a,c}$ represents the number of susceptible to SARS-CoV-2 infection in the population class *{a,c}*, where *a* represents the age group and *c* identifies the absence/presence of underlying conditions.

$I_{a,c}$represents the number of infectious unvaccinated individuals in the population class *{a,c}.*

$R_{a,c}$represents the number of unvaccinated individuals in the population class *{a,c}* who recovered from infection.

$V_{0,a,c},V_{1,a,c},{and V}_{2,a,c}$ represent the number of vaccinated individuals in each ramp-up stage. In particular,

1) $V_{0,a,c}$denotes individuals in the population class *{a,c}* vaccinated with the first dose. In the main analysis, we assumed that the second dose is administered 21 days after the 1^st^ dose. So ${1/\omega}_{0}=21$ days.

2) $V_{1,a,c}$ denotes individuals in the population class *{a,c}* vaccinated with the second dose, for whom the 2^nd^ dose is not effective yet. We assumed that the second dose becomes effective 14 days after administration, so ${1/\omega}_{1}=14$ days.

3) $V_{2,a,c}$ denotes individuals in the population class *{a,c}* vaccinated with the second dose for whom vaccination is effective.

$I_{a,c}^{\mathrm{vaccinated}}$represents the number of infectious individuals in the population class *{a,c}* among those who have already received at least one dose of vaccination.

$R_{a,c}^{\mathrm{vaccinated}}$represents the number of individuals in the population class *{a,c}* who developed infection despite having received vaccination (one or more doses).

Susceptible individuals are exposed to a time and age-dependent force of infection$\lambda_{a}\left( t \right)$ which is defined as:

$$\lambda_{a}\left( t \right)=\left( 1-\varphi\right)\beta r_{a}\sum_{\tilde{a}} C_{a,\tilde{a}}\frac{\sum_{c} [I_{\tilde{a},c} \left( t \right)+I_{\tilde{a},c}^{vaccinated}\left( t \right)]}{\sum_{c} N_{\tilde{a},c}}$$

where:

$\beta$ is a scaling factor shaping SARS-CoV-2 transmissibility in the absence of non-pharmaceutical interventions (no NPIs, Effective reproductive number R_t_ =2.5), such as social distancing, school closure, and case isolation.

$\varphi$ is a coefficient representing the reduction in transmissibility due to NPIs.

$r_{a}$is the relative susceptibility to SARS-CoV-2 infection at age $a$: $r_{a}=0.58$ (95%CI 0.34-0.98) when $a<15$; $r_{a}=1$ for $15\leq a<65$*;* $r_{a}=1.65$ (95%CI 1.03-2.65) when $a\geq65$ ^4,5^.

$C_{a,\tilde{a}}$ represents the age-group-specific contact matrix, whose entries describe the mean numbers of persons in age group $\tilde{a}$ encountered by an individual of age group $a$ per day.

$N_{\tilde{a},c}$ represents the number of individuals in the population class *{*$\tilde{a}$*,c}*.

For all infectious compartments, the average duration of infectiousness $(1/\gamma)$ is set equal to the average generation time (5.5 days)^4^.

At each time *t*, the first dose of vaccination is administered to a fraction $\alpha_{a,c}\left( t \right)$of susceptible individuals in the population class *{a,c}*:

$$\alpha_{a,c}\left( t \right)=\frac{d_{a,c}(t)}{S_{a,c}\left( t \right)}$$

where $d_{a,c}(t)$ represents the number of (first) vaccine doses to be administered to individuals of the population class *{a,c}* at time *t* under the considered vaccination scenario.

The daily number of first doses $d_{a,c}\left( t \right)$ to be administered to the population class *{a,c}* is computed by taking into account: i) the assumed priority order; ii) the assumed vaccination coverage, i.e. the fraction of population that is expected to be vaccinated at the end of the program; ii) the constraints on the daily vaccination capacity. In particular, we assume that half of the daily capacity is allocated to first doses, i.e.:

$\sum_{a,c} d_{a,c}\left( t \right)$= (daily vaccination capacity)/2

and the remaining half to second doses.

Vaccinated individuals $V_{i,a,c} \left( \text{i=0,1,2} \right)$can develop infection, but their susceptibility to infection is reduced by a factor ($1-{VE}_{i,a})$,where ${VE}_{i,a}$represents the age-specific vaccine efficacy associated to the *i*-th vaccination stage. In the main analysis, the age-dependent vaccine efficacy after the 1^st^ dose of vaccination (${VE}_{0,a})$was assumed to be 0; the age-dependent efficacy right after administration of the 2^nd^ dose (${VE}_{1,a})$was also assumed to be 0; while the vaccine efficacy after ramp-up of the 2^nd^ dose (${VE}_{2,a})$ was assumed to be 80% for individuals aged 20-59 years and 40% for all other age groups. Simulation results discussed in the main text and in the following sections were obtained by using a stochastic version of the model described above.

A summary of model parameters and data sources is presented in Table 1.

**Table 1. Description of key parameters used in the model.**

| **Description of parameter** | **Values in the baseline analysis** | **Sensitivity analyses (SA)** |
| --- | --- | --- |
| **Epidemiology** |  |  |
| Generation time (1/$\gamma)$ | 5.5 days (95%CI 1.7, 11.6) ^*4^ | / |
| Relative susceptibility to infection at age $a$ ($r_{a}$) | $r_{a}=0.58$ (95%CI 0.34-0.98) when $a<15$; $r_{a}=1$ for $15\leq a<65$; $r_{a}=1.65$ (95%CI 1.03-2.65) when $a\geq65$ ^4^ | Homogenous susceptibility *(****SA19****)* |
| Age-group-specific contact matrix ($C_{a,\tilde{a}}$) | Contact matrix for Shanghai before pandemic ^1^ | Contact matrix for Shanghai at the last stage of the first wave of pandemic in Wuhan, China (March 2020) *(****SA13****)^6^* |
| Effective reproductive number (R_t_) | 1.1, 1.3, 1.5, and 2.5 ^7-12^ | / |
| **Vaccination** |  |  |
| Interval between the administration of 1^st^ dose and 2^nd^ dose (1/$\omega_{0}$) | 21 days^*^ ^13^ | 14 and 28 days *(****SA16-SA17****)* ^13^ |
| Delay between administration of the 2^nd^ dose of vaccination and the achievement of the expected VE (1/$\omega_{1}$) | 14 days^*^ ^13^ | / |
| Expected vaccine efficacy for adults aged 20-59 years (${VE}_{2,a}$) | 80% ^13,14^ for a vaccine with partial protections | 60% *(****SA9****)* and 90% *(****SA10****);* and for an all-or-nothing vaccine *(****SA18****)* |
| Expected vaccine efficacy reduction for <20 and ≥60 years | 50% ^13,14^ | 0%, indicating the same vaccine efficacy *(****SA24****)* |
| Vaccination capacity (daily doses administered) | 6 million (Assumed based on 2009 influenza pandemic vaccination) ^15^ | 1.3 *(****SA20****)*, 10 *(****SA21****)*, 15 *(****SA22****)* and 30 million *(****SA23****)* |
| Vaccine coverage | Homogenous across age groups: 70% ^3^ | Homogenous across age groups: 50% (***SA4***) or 90% *(****SA5****) ^3^;* 70% for adults≥20 years, and then 50% for others (***SA6***); 90% for adults ≥20 years, and then 70% for others (***SA7***); 70% for adults ≥20 years, and then no vaccination for others (***SA8***) |
| Duration of immunity (1/$\omega_{2}$) | Lifelong (i.e., the immunity lasts more than the time horizon considered: 730 days)  (Assumed) | 6 months *(****SA12****)*, or 1 year *(****SA25****)* ^*^ |
| Delay between start of simulations and start of vaccination | 15 days (Assumed) | SARS-CoV-2 infections leading to an outbreak are imported when 10% (***SA1***), 20% (***SA2***), and 30% (***SA3***) of the Chinese population has already been vaccinated. |
| **Disease burden** |  |  |
| Proportion of infections that develop symptoms ($\pi$) | 18.1%, 22.4%, 30.5%, 35.5%, and 64.6% separately for 0-19, 20-39, 40-59, 60-79, and 80+ years^16^ | / |
| Proportion of laboratory-confirmed symptomatic cases requiring hospitalization ($\sigma$) | Overall: 40.0%, 29.2%, 33.3%, and 33.8% separately for 0-19, 20-39, 40-59, and 60+ years^17^; estimates for individuals with/without underlying conditions shown in Supplementary Information File 4 | / |
| Proportion of hospitalized cases requiring ICU ($\rho$) | 0, 2.2%, 7.2%, 20.9% separately for 0-14, 15-49, 50-64, and 65+ years^18^ | / |
| Fatality ratio among laboratory-confirmed symptomatic cases ($\mu$) | Overall: 0.51%, 0.65%, 2.38%, and 10.52% separately for 0-19, 20-39, 40-59, and 60+ years^17^; estimates for individuals with/without underlying conditions shown in Supplementary Information File 4 | / |

*mean value.

### Supplementary file 2. Estimating of the scaling factor for transmissibility in the absence of NPIs ($\boldsymbol{\beta}$)

The reproduction number can be computed as the dominant eigenvalue of the Next Generation Matrix (NGM) ^19^ associated with the dynamical system considered:

$${(NGM)}_{a,\tilde{a}}=\frac{\beta}{\gamma}r_{a}C_{a,\tilde{a}}$$

We assumed a reproduction number in the absence of NPIs $R_{t}\left( \text{no}\text{ }\text{NPIs} \right)=2.5$ (4,8,9). Given the value of $R_{t}\left( \text{no}\text{ }\text{NPIs} \right)$, the distribution of the age-specific susceptibility profile ($r_{a})$and the distribution of the bootstrapped contact matrix, we computed the distribution of $\beta$ analytically.

When considering a set of NPIs that are capable to bring the reproduction number to a value $R_{t}(\text{NPIs})$< $R_{t}(\text{no}\text{ }\text{NPIs})$, we used the distribution of $\beta$ obtained in the absence on NPIs, rescaled by a factor ($1-\varphi)$ where

$\varphi=1-R_{t}(\text{NPIs})$/$R_{t}(\text{no}\text{ }\text{NPIs})$.

### Supplementary file 3. COVID-19 vaccine doses distributed over time in China

The Joint Prevention and Control Mechanism of the State Council in China released a three-step strategy for COVID-19 vaccine rollout ^20^. We systematically collected information about COVID-19 vaccination in China from the website of the State Council of the People’s Republic of China ^16^. The number of daily doses administered is less than 3 million in the first stage and shows a growing trend from 3 to 10 million, with a daily average of about 6 million doses over the period April 20 – May 7, 2017. As of May 7, a total of 308 million doses have been administered.

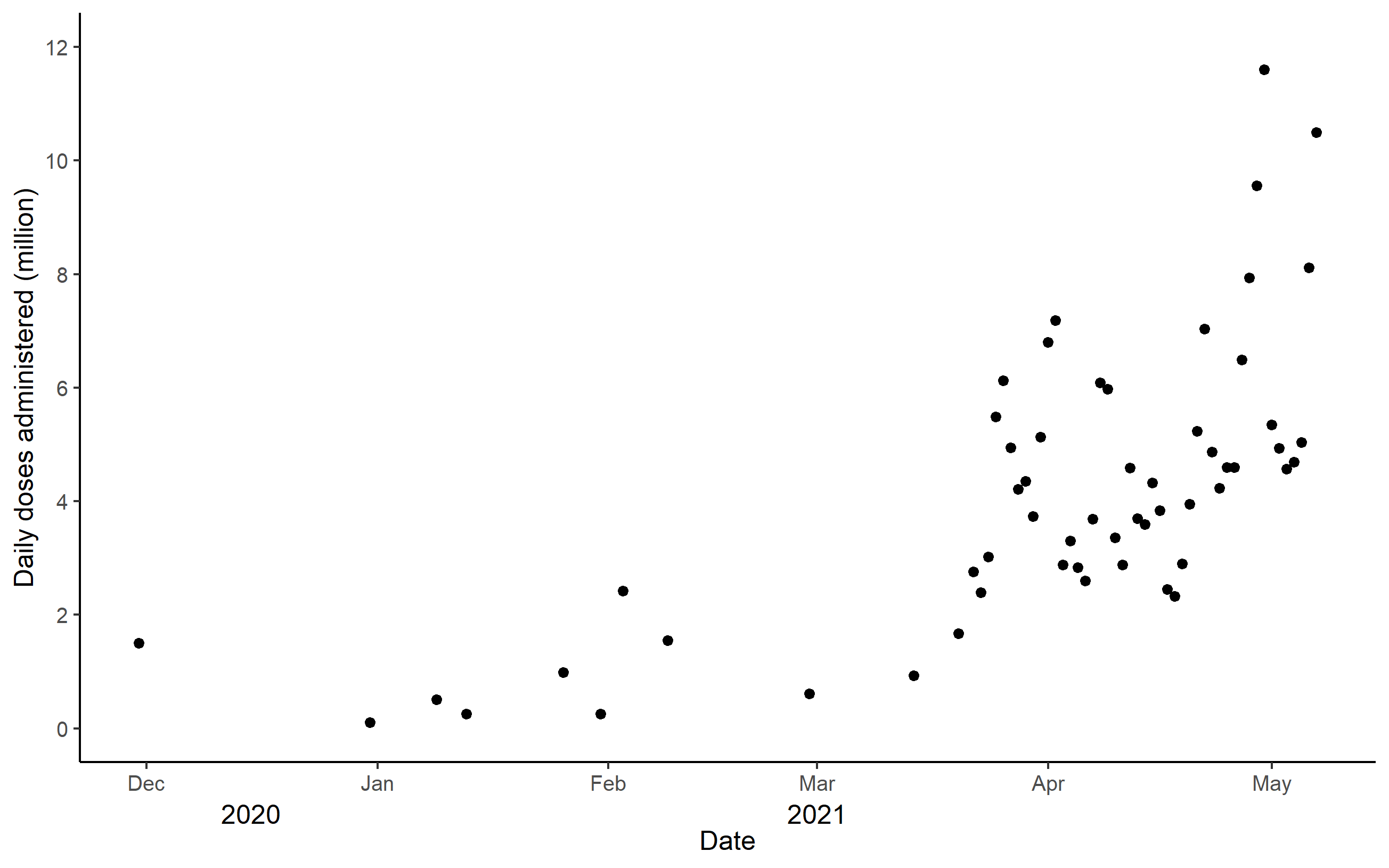

Fig. 2 Doses of COVID-19 vaccines administered per day in China, as of May 7, 2021

### Supplementary file 4. Priority population of COVID-19 vaccination

**Table 2. Priority population of COVID-19 vaccination***

| Tier of vaccination | Baseline (First prioritization to old adults and individuals with underlying conditions) | First prioritization to old adults  (***SA26^ǂ^***) | First prioritization to working-age groups (***SA27***) | First prioritization to school-age groups (***SA28***) |
| --- | --- | --- | --- | --- |
| 1 | Healthcare workers (No=10.7 million) | | | |
| 2 | Law enforcement and security workers, personnel in nursing home and social welfare institutes, community workers, workers in energy, food and transportation sectors, etc.  (No=36.8 million) | | | |
| 3 | Adults ≥ 60 years of age with underlying conditions, and adults ≥ 80 years of age without underlying conditions  (No.=162.9 million) | Adults ≥ 60 years of age  (No= 248.6 million) | Individuals aged 20-59 years  (No= 807.2 million) | School-age children  (No= 237.4 million) |
| 4 | Older adults aged 60-79 years without underlying conditions, individuals aged < 60 years with pre-existing medical conditions, and pregnant women  (No.=401.2 million) | Individuals aged 20-59 years  (No= 807.2 million) | School-age children  (No= 237.4 million) | Individuals aged 20-59 years  (No= 807.2 million) |
| 5 | Individuals aged 20-59 years without underlying conditions  (No.=525.8 million) | School-age children  (No= 237.4 million) | Adults ≥ 60 years of age  (No= 248.6 million) | Adults ≥ 60 years of age  (No= 248.6 million) |
| 6 | School-age children and younger children ≤5 years  (No.=301.9 million) | Younger children≤5 years  (No= 98.7 million) | | |

**^*^**Healthcare workers and the other essential workers listed here are fixed in Tier 1 and Tier 2 of vaccination, and thus would be vaccinated before other subgroups. ^ǂ^Sensitivity analysis.

### Supplementary file 5. Estimating the proportion of laboratory-confirmed COVID-19 symptomatic cases requiring hospitalization and death for individuals with and without underlying conditions

In order to quantify the different burden of COVID-19 in individuals with and without underlying conditions (such as chronic respiratory disease, heart disease, cardio-cerebrovascular disease, hypertension, diabetes, chronic renal diseases, chronic liver disease, cancer, and obesity ^3^), we estimated the hospitalization and death rates for the two subgroups in China, using below data: 1) the overall age-specific hospitalization and death rates among symptomatic cases independent from the presence of underlying conditions in China ^17^; 2) the proportion of symptomatic cases hospitalized/died in the two subgroups as obtained from the Lombardy region of Italy ^16,21^.

The age-specific proportions of laboratory-confirmed symptomatic cases requiring hospitalization for individuals with ($\sigma_{a,u}$) and without ($\sigma_{a,nu}$) underlying conditions were computed respectively as:

$\sigma_{a,u} =s\cdot h_{u}^{ITA}\cdot$ $\Delta_{a}$

$\sigma_{a,nu} =s\cdot h_{nu}^{ITA}\cdot$ $\Delta_{a}$

Where,

- $h_{u}^{ITA}$ and$h_{nu}^{ITA}$ separately denote the proportion of hospitalized among symptomatic cases with and without underlying conditions as estimated from Lombardy data (Table 2) ^16,21,22^.
- $\Delta_{\mathbf{a}}$ denotes the age-specific proportion of laboratory-confirmed symptomatic cases requiring hospitalization as estimated for China independently from the presence of underlying conditions ^17^.
- the scale factor $s$ is determined in such a way to minimize the root mean square error between $\Delta_{a}$ and $\tilde{\Delta_{a}}=$ $P_{a,u}\cdot\sigma_{a,u} +P_{a,nu}\cdot\text{ }\sigma_{a,nu}.$ $P_{a,u}$and $P_{a,nu}$ denote the proportions of individuals of age with and without underlying conditions in China, respectively ^3^.

Analogously, the age-specific fatality ratios among laboratory-confirmed symptomatic cases for individuals with ($\mu_{a,u}$) and without ($\mu_{a,nu}$) underlying conditions are computed respectively as:

$\mu_{a,u} =v\cdot m_{u}^{ITA}\cdot$ $M_{a}$

$\mu_{a,nu} =v\cdot m_{nu}^{ITA}\cdot$ $M_{a}$

where,

- $m_{u}^{ITA}$ and$m_{nu}^{ITA}$ denotes the proportion of cases with fatal outcomes among symptomatic cases with and without underlying conditions as estimated from Lombardy data (Table 3) ^16,21,22^.
- $M_{a}$ denotes the age-specific fatality ratio among laboratory-confirmed symptomatic cases as estimated for China independently from the presence of underlying conditions ^17^.
- the scale factor $v$ is determined in such a way to minimize the root mean square error between $M_{a}$ and $\tilde{M_{a}}=$ $P_{a,u}\cdot\mu_{a,u} +P_{a,nu}\cdot\text{ }\mu_{a,nu}$.

Estimates were reported in Table 4.

**Table 3. Proportion of laboratory-confirmed symptomatic cases requiring hospitalizations and having fatal outcomes among patients with or without underlying conditions***

|  | With underlying conditions | Without underlying conditions | Total |
| --- | --- | --- | --- |
| Laboratory-confirmed symptomatic cases | 44446 | 44092 | 88538 |
| Laboratory-confirmed symptomatic cases requiring hospitalizations | 29593 | 17800 | 47393 |
| Laboratory-confirmed symptomatic cases with fatal outcomes | 13683 | 3095 | 16778 |
| Proportion of hospitalization among laboratory-confirmed symptomatic cases (%) | $h_{u}^{ITA}=$66.6 | $h_{nu}^{ITA}=$40.4 | 53.5 |
| Proportion of laboratory-confirmed symptomatic cases with fatal outcomes (%) | $m_{u}^{ITA}=$30.8 | $m_{nu}^{ITA}=$7 | 19 |

^*^ The data were obtained from the line list of COVID-19 patients in the Lombardy region of Italy, with underlying diseases including chronic respiratory disease, cardiovascular disease, metabolic disease and cancer ^16,21,22^.

**Table 4. Estimated hospitalization and death rates for individuals with and without underlying conditions in China.**

|  | With/without underlying conditions | With underlying conditions | Without underlying conditions |
| --- | --- | --- | --- |
| Proportion of laboratory-confirmed symptomatic cases requiring hospitalizations (%) | $\Delta_{a}$^17^ | $\sigma_{a,u}$ | $\sigma_{a,nu}$ |
| 0-19 years | 40 | 51.9 | 31.5 |
| 20-39 years | 29.2 | 37.9 | 23.0 |
| 40-59 years | 33.3 | 43.2 | 26.2 |
| 60+ years | 33.8 | 43.8 | 26.6 |
| Fatality ratio among laboratory-confirmed symptomatic cases (%) | $M_{a}$^17^ | $\mu_{a,u}$ | $\mu_{a,nu}$ |
| 0-19 years | 0.51 | 0.66 | 0.15 |
| 20-39 years | 0.65 | 0.84 | 0.19 |
| 40-59 years | 2.38 | 3.06 | 0.70 |
| 60+ years | 10.52 | 13.53 | 3.07 |

### Supplementary file 6. Data analysis

For each scenario, 200 stochastic model realizations were performed. The outcome of these simulations determined the distributions of the number of symptomatic infections, hospitalizations, ICU admissions, and deaths. 95% confidence intervals were defined as quantiles 0.025 and 0.975 of the estimated distributions. We used a Bayesian approach to estimate R_t_ from the time series of symptomatic cases by date of symptom onset and the distribution of the serial interval. The methods have been described previously ^23^.

To estimate R_t_, we assumed that the daily number of new cases (by date of symptom onset), including locally acquired infections L(t), can be approximated by a Poisson distribution according to the equation.

$$L(t)\sim Pois\left( R(t)\sum_{s=1}^{t} \varphi\left( s \right)C(t-s) \right)$$

Where,

- $C(t)$, with t from 0 to T, is the daily number of locally acquired new cases, by date of symptom onset;
- $R(t)$ is the net reproduction number at time t;
- $\varphi(s)$ is the distribution of the generation time (corresponding to the distribution of the serial interval) calculated at time s.

The likelihood ℒ of the observed time series of cases from day 1 to T conditional on $C(0)$ is thus given by

$$\mathcal{L=}\prod_{t=1}^{T} P\left( L\left( t \right);R(t)\sum_{s=1}^{t} \varphi\left( s \right)C(t-s) \right)$$

where $P(k; \lambda)$ is the probability mass function of a Poisson distribution (i.e., the probability of observing k events if these events occur with rate λ).

We used Metropolis-Hastings MCMC sampling to estimate the posterior distribution of $R(t)$. The Markov chains were run for 1,000,000 iterations, assuming non-informative prior distributions of $R(t)$ (flat distribution in the range (0-1000]). Convergence was checked by visual inspection by running multiple chains starting from different starting points. It should be noted that, when estimating $R(t)$, we excluded data at the start and end of the epidemic curve for these days with the first 5% quantile of daily number of symptomatic cases to deal with possible instability of $R(t)$ estimates due to small numbers of symptomatic cases. While for $R(t)$=1.1 at the beginning of transmission, we do not exclude aforementioned data since the daily number of symptomatic cases remains tiny during the time frame of this study.
